# Biological age measured by DNA methylation clocks and mortality: A systematic review and meta-analysis

**DOI:** 10.64898/2026.10.20.26365864

**Authors:** Jian Hua Tay, Duarte Barros, Jiajun Dong, Shubham Choudhury, Austeja Bakulaite, Weilan Wang, Andrea B. Maier

**Author notes:** These authors share first authorship and contributed equally to this work (Jian Hua Tay, Duarte Barros, Jiajun Dong). Corresponding author: Prof Andrea B. Maier, Department of Human Movement Sciences, @AgeAmsterdam, Faculty of Behavioural and Movement Sciences, Amsterdam Movement Sciences, Vrije Universiteit, Van Der Boechorstsraat 7, 1081 BT Amsterdam, The Netherlands. Linkedin: https://www.linkedin.com/in/andreamaierprof/.

## Abstract

This systematic review aimed to synthesize evidence on the association between DNA methylation (DNAm) age and all-cause and disease-specific mortality risks in adults. A systematic search of six databases from Jan 1 2011 to April 15 2026 identified articles assessing the association of DNAm age measures and all-cause or disease-specific mortality in adults from general or disease-specific populations. Effect sizes were pooled as hazard ratios (HRs) with 95% confidence intervals (CIs) using inverse-variance random-effects models. Of 42 023 records screened, 125 articles (380 705 participants) met the inclusion criteria, with 53 included in meta-analyses. Overall, 48 distinct DNAm clocks were evaluated. Higher DNAm ages were associated with higher risks of all-cause, cardiovascular, and cancer mortality risks. In general populations, GrimAge epigenetic age acceleration (EAA) showed consistent associations across mortality outcomes, with each 1 year higher GrimAge EAA associated with ∼6-10% higher all-cause (31 cohorts, n=53 642, HR per SD: 1.46 [95% CI 1.38, 1.55], HR per 1-year: 1.09 [95% CI 1.08, 1.11], I²=84.1%, p-value=1.21×10⁻¹⁴), cardiovascular (6 cohorts, n=30 751, HR per SD: 1.55 [95% CI 1.38, 1.73], HR per 1-year: 1.10 [95% CI 1.07, 1.13], I²=28.0%, p-value=1.71×10⁻⁴), and cancer (5 cohorts, n=27 187, HR per SD: 1.31 [95% CI 1.15, 1.50], HR per 1-year: 1.06 [95% CI 1.03, 1.10], I²=33.6%, p-value=4.76×10⁻³) mortality. Clocks incorporating additional surrogates and technical refinements showed numerically larger pooled associations than their original counterparts. GrimAge2 (HR per SD: 1.65 [95% CI 1.53, 1.77] vs 1.46 [1.38, 1.55]) and PCGrimAge (1.56 [1.36, 1.78] vs 1.51 [1.32, 1.74]) showed higher pooled HR association with all-cause mortality than GrimAge. Further work is warranted to evaluate the responsiveness of DNAm clocks to interventions and potential clinical utility.

## Introduction

The leading causes of mortality projected until 2050 are shifting towards to age-related diseases, with the leading four causes being ischaemic heart disease, stroke, diabetes, and chronic obstructive pulmonary disease.^1^ Biological age is a stronger predictor of mortality and disease risk than chronological age,^2^ potentially allowing early identification of individuals at elevated risk before clinical decline and guiding geroprotective interventions.^3^ Deoxyribonucleic acid (DNA) methylation (DNAm)-based clocks, also known as epigenetic clocks, are potential candidates, as they can be reliably measured from blood, and demonstrate high test–retest technical reliability.^3^ Several generations of DNAm clocks were developed, from predicting chronological age,^4,5^ mortality risk,^2,6,7^ pace of aging,^8,9^ to biological mechanism and interpretability.^10,11^

Meta-analysis of four cohort studies found that Horvath epigenetic age acceleration (EAA) was associated with a 16% higher 5-year all-cause mortality risk.^12^ GrimAge and GrimAge2 outperformed first-generation clocks, Horvath and Hannum, and were associated with 44% and 40% higher 10-year all-cause mortality risk with 1-year higher EAA, respectively.^13^ The previous systematic reviews and meta-analyses of DNAm age and mortality have been limited to Horvath, Hannum, PhenoAge and GrimAge. These reviews did not include clocks established 2022 onwards, such as DunedinPACE, GrimAge2 and bAge, that may have stronger mortality prediction ability.^14–16^ As DNAm clocks move closer to clinical and risk-stratification application, an updated synthesis of evidence is warranted to assess the performance of DNAm age as a surrogate biomarker for mortality, and identify the DNAm clocks that best predict mortality.^3^ Analysing their predictive ability across all-cause and disease-specific mortality is important for regulatory assessment and potential implementation.^3^

This systematic review aimed to assess the predictive ability of DNAm age for all-cause or disease-specific mortality in general and specific disease population.

## Method

### Search strategy and selection criteria

This systematic review followed the PRISMA guidelines.^17^ A comprehensive search was performed across six electronic databases, including Embase, Cochrane Central Register of Controlled Trials (CENTRAL), PubMed, Ovid (PsycINFO and Medline), Scopus, and Web of Science, using MeSH terms and keywords related to DNA methylation clocks (**Supplementary Text 1 and Supplementary Table 1**). The search included articles published between Jan 1, 2011, and April 15, 2026, as the first DNA methylation clock was published in 2011.^18^ Search terms were developed with support from a senior librarian (Loh Mee Lan, National University of Singapore). Additional articles were identified through the reference lists of included articles. The protocol was registered in PROSPERO (**CRD42024499021**). Methodological details are provided in prior publications.^19^

### Article selection

Two assessors (JHT and WW) independently removed duplicates and then screened titles and abstracts for eligibility using Covidence systematic review software (Veritas Health Innovation). Full-text articles were screened independently (by JHT and WW) and categorised into specific age-related diseases, conditions, and health outcomes using the tagging function of Covidence. Other epigenetic clocks not based on DNAm, including assay for transposase-accessible chromatin (ATAC) clock^20^ and the histone clock,^21^ were excluded. Discrepancies were resolved by a third author (ABM). Peer reviewed articles were included if they reported on population-based cohort studies published in English, including adults aged 18 years or older from general populations or disease-specific populations. Eligible studies assessing the predictive ability of DNA methylation ages for mortality (all-cause and disease-specific mortality) from minimally invasive sample collection methods (including blood, saliva, buccal cell, or skin samples) were considered. Review articles, studies without primary data and studies not reporting on DNAm clocks were excluded. Only articles that included DNAm age metrics, including DNAm age, DNAm age acceleration (epigenetic age acceleration (EAA) or relative age acceleration), DNAm age deviation (AgeDev or absolute age acceleration) measurements, or pace of ageing, were considered.^3,22^ Detailed inclusion criteria and definitions are provided (**Supplementary Text 1)**.

The DNAm clocks were categorised into four generations. First-generation DNAm clocks have been trained on chronological age including Horvath1,^4^ Hannum,^5^ Horvath2,^23^ Lin,^24^ Weider,^25^ Vidal-Bralo,^26^ Zhang2019,^27^ 7-CpG,^28^ 27CpGWKDE,^29^ second-generation clock trained on health outcomes including PhenoAge,^2^ GrimAge,^6^ GrimAge2,^7^ ZhangMRscore,^30^ DNAmFitAge,^31^ 59-CpG,^6^ MetabolicAge,^32^ MetaboHealth,^33^ DNAm IC,^34^ aSES-BIO, ^35^ cSES-BIO,^35^ bAge,^36^ CheekAge,^37^ InflammAge,^38^ DNAmTL,^39^ EpiTOC,^40^ third-generation clocks trained on pace of ageing with longitudinal health outcomes including DunedinPoAm^8^ and DunedinPACE,^9^ fourth generation clocks based on biological interpretability including causality enriched clocks^10^ and System Age. ^11^

### Data extraction and quality assessment

Pairs of assessors (JHT, DB, JD, SC, AB) independently extracted data using a standardised form adapted from previous systematic reviews,^14–16^ including title, first author, publication year, study design, cohort location, participants’ characteristics (sample size, group size, mean or median age, age range, and sex), DNAm source, DNAm profiling method (including whole-genome bisulfite sequencing, Illumina array, or other technology), DNAm clock used, effect estimates (odd ratio (OR), hazard ratio (HR), relative risk ratio (RR)), and adjustment models applied. Study quality was independently assessed by two reviewers (JHT and DB) using the Newcastle–Ottawa Scale for cohort studies (**Supplementary Text 2**).^41^ Discrepancies were resolved by a third author (ABM).

### Data synthesis and meta-analysis

Heatmaps were created to illustrate the association between DNAm based clocks and all-cause and disease-specific mortality across all included studies, separately for the general population and disease-specific populations. Estimates were based on fully adjusted models.

Meta-analyses were conducted for each DNAm clock with at least three independent cohorts reporting HRs and 95% confidence intervals (CIs) for all-cause or disease-specific mortality, in populations with similar baseline characteristics. For cohorts represented by multiple publications, a single estimate was included, selecting the study with the greatest sample size and/or longest follow-up.^40^ General-population and disease-specific cohorts were analysed separately. Disease-specific cohorts were evaluated for clinical heterogeneity to determine for suitability of meta-analyses. Clinical heterogeneity refers to variability between studies in the characteristics of the study populations that may influence effect estimates.^42^

For clinical interpretability, all HRs were harmonised to represent the risk associated with a 1-year higher in EAA, a 0.1-unit higher in MRscore or a 0.1-unit higher in pace of ageing, where possible. Otherwise, HRs were reported per one standard deviation increase in EAA. For studies that did not report HR per unit higher in EAA, exponential rescaling was performed to estimate the HR for unit higher EAA. For example, if a study reported an HR of 1.20 per 5-year higher in EAA, this was converted to an HR of approximately 1.04 per 1-year higher in EAA. To aid interpretation, the HR represents the relative rate of mortality during follow-up. For example, a HR of 1.5 indicates that each 1-year (or 0.1-unit or 1 SD, depending on the clocks) higher EAA is associated with a 50% higher hazard of mortality. Main analyses were based on minimally adjusted models (e.g. age and sex) and fully adjusted models with age, sex, cell composition, lifestyle and other factors as covariates, with the most comparable estimates selected when multiple adjustment sets were reported. Secondary analyses assessed whether Principal component (PC) clocks and technical refinement of existing clocks with added biomarkers (e.g. GrimAge to bAge) improved clock performance. PC-based clocks included PCHorvath1, PCHorvath2, PCHannum, PCPhenoAge, PCGrimAge, and PCDNAmTL, which were derived by applying principal component analysis (PCA) to the CpG components of the original Horvath1, Horvath2, Hannum, PhenoAge, GrimAge, and DNAmTL clocks to enhance reliability and reduce technical noise.^43^ Other technical refinement includes enhancement of existing clocks with additional DNAm proxies, for instance, bAge is refined from GrimAge with additional protein proxies.^36^ Secondary analyses were conducted within the same cohorts using identical models to ensure direct comparability.

Prior to meta-analysis, HRs from individual studies were log-transformed. Pooled log-transformed HRs were synthesized using inverse-variance weighted random effect meta-analysis with Hartung-Knapp adjustment, then back-transformed (exponentiated) to HRs for data presentation and forest plot visualization.^42^ For comparability, pooled HRs per SD were compared across clocks to facilitate direct comparison of predictive ability, with assistant of the p values from meta-analyses (p-values) since clocks were reported on different scales (e.g., 1-year higher for year-based clocks, and 0.1-unit higher for the pace of ageing).^6,7^ Between-study heterogeneity was quantified using the I² statistic, Cochran’s Q (χ² test), and τ² estimates.^42^ All analyses were performed in R (version 4.6.1) using the metafor packages. Funnel plots and Egger’s regression test were used to assess small-study effects and potential publication bias when there were at least ten studies.^42^ Sensitivity analyses were conducted to evaluate the robustness of the findings: (1) repeating meta-analyses with only high quality studies where available and (2) repeating meta-analyses excluding studies from which conversion of HRs relied on imputed standard deviation. Further details are provided (**Supplementary Text 1**).

## Results

### Study characteristics

A total of 42 023 records were identified in the literature search, 24 896 duplicates were removed, 14 718 were excluded based on titles and abstracts. Out of 2409 full-text articles assessed for eligibility, 719 excluded due to ineligibility, 1565 were excluded as they investigated health outcomes unrelated to mortality, 125 met the eligibility criteria and were included in the systematic review (**Figure 1**).

**Fig. 1:**
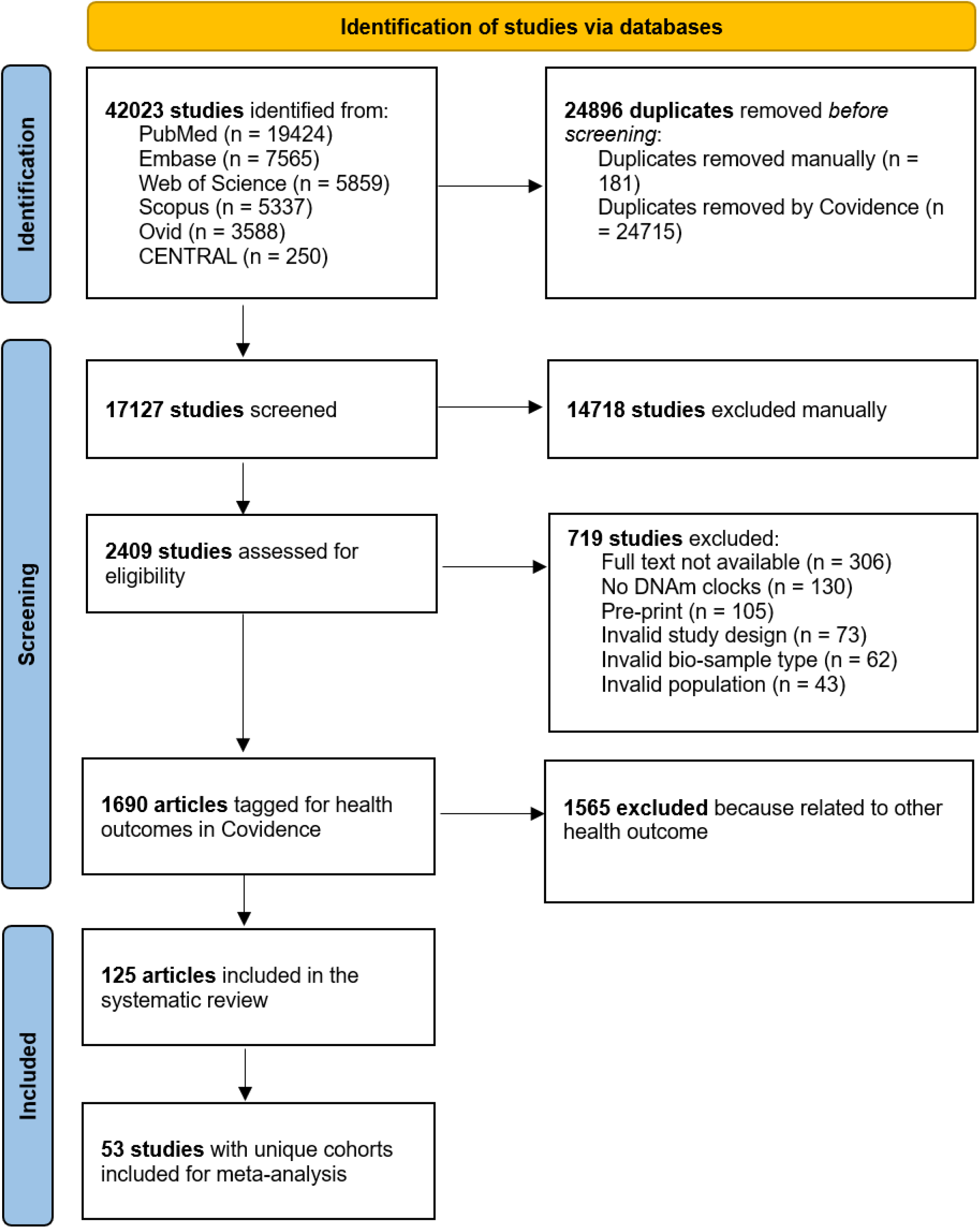
Flow chart of search strategy.

The 125 articles included 380 705 participants (∼137 000 unique participants). Of these, 96 were conducted in individual cohorts, while 29 included analyses across multiple cohorts. Individual study sample sizes ranged from 12 to 18 869 participants. The following cohorts were frequently reported: Framingham Heart Study (FHS, n=18)^2,6,7,9,10,12,34,36,39,44–52^, The Health and Retirement Study (HRS, n=17)^32,35,53–67^, National Health and Nutrition Examination Survey (NHANES, n=17)^13,55,68–82^, WHI (n=11)^2,6,7,11,23,31,36,39,45,51,83^, The Normative Aging Study (NAS, n=11)^2,7–10,12,45,84–87^, or Jackson Heart Study (JHS, n=7)^2,6,7,18,23,31,39,51^ located in the USA; and the Lothian Birth Cohorts 1921 (LBC1921, n=11)^7,12,24,27,29,31,36,37,39,45,88^ and 1936 (LBC1936, n=12)^7,12,24,27,29,31,36,37,39,45,88,89^, and The Invecchiare in Chianti (InCHIANTI, n=7)^6,7,23,31,39,45,90^ cohorts in Europe. The follow-up time of studies ranged from 0.25 to 30 years. Most articles were conducted in the general population (n=96),^2,6–13,23,24,27,29–39,44–66,68–70,72,73,76–120^ with 52 articles including cohorts with a mean or median age ≥65 years (**Table 1**). One article reported data on general population and cancer-survivors,^67^ while 28 articles focused on disease-specific populations, including trauma-exposed veterans,^121^ cancer survivors,^122,123^ patients with cancer,^124–132^ schizophrenia,^133^ amyotrophic lateral sclerosis,^134^ human immunodeficiency virus infection,^135–138^ acute ischemic stroke,^139^ pre-diabetes or type 2 diabetes,^74,140,141^ kidney disease,^71,142^ geriatric population with fractures^143^, rheumatoid arthritis^75^, patients with amyotrophic lateral sclerosis^144^ or individuals undergoing cardiac catheterisation^145^ (**Supplementary Table 2**).

**Table 1.**
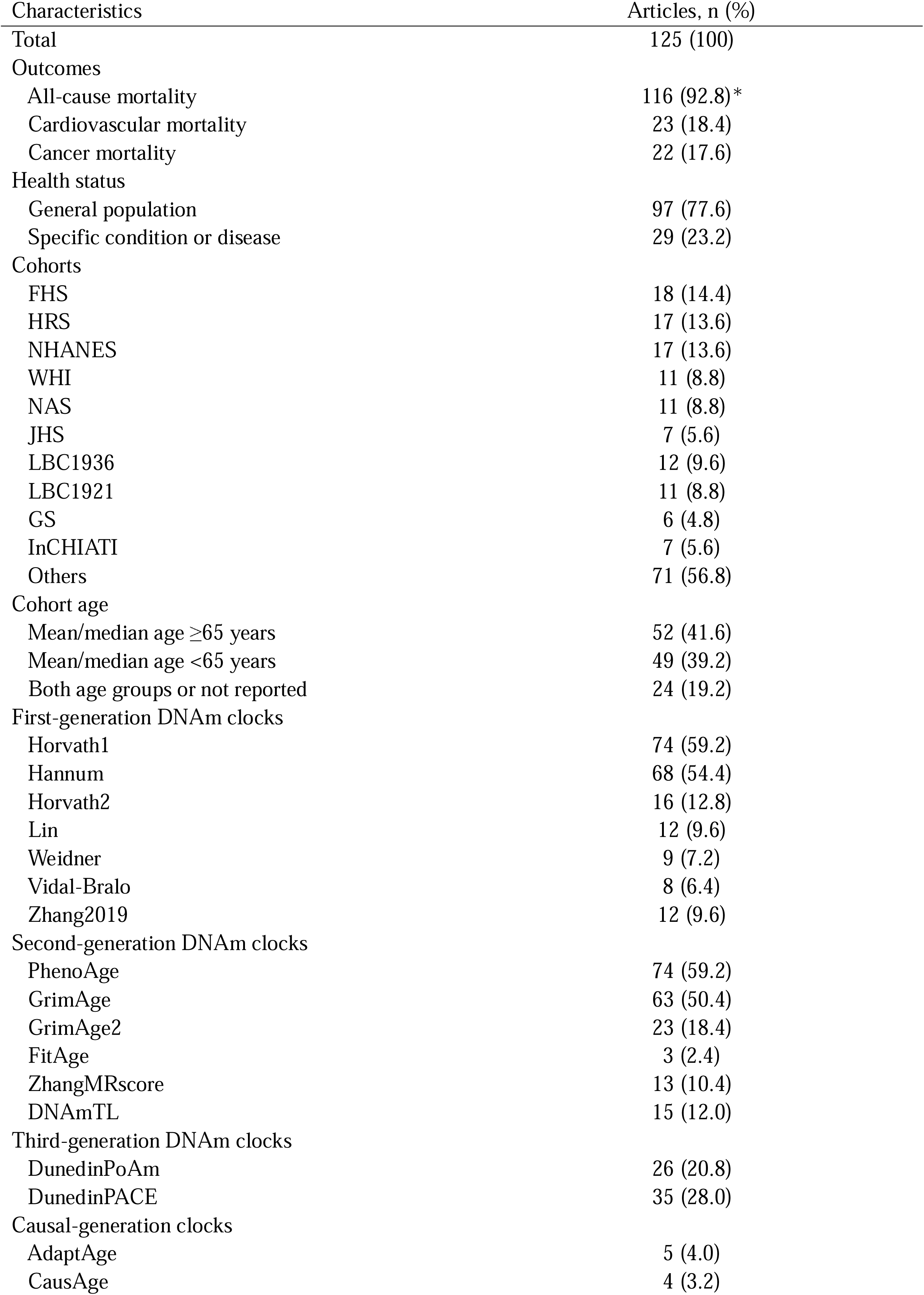
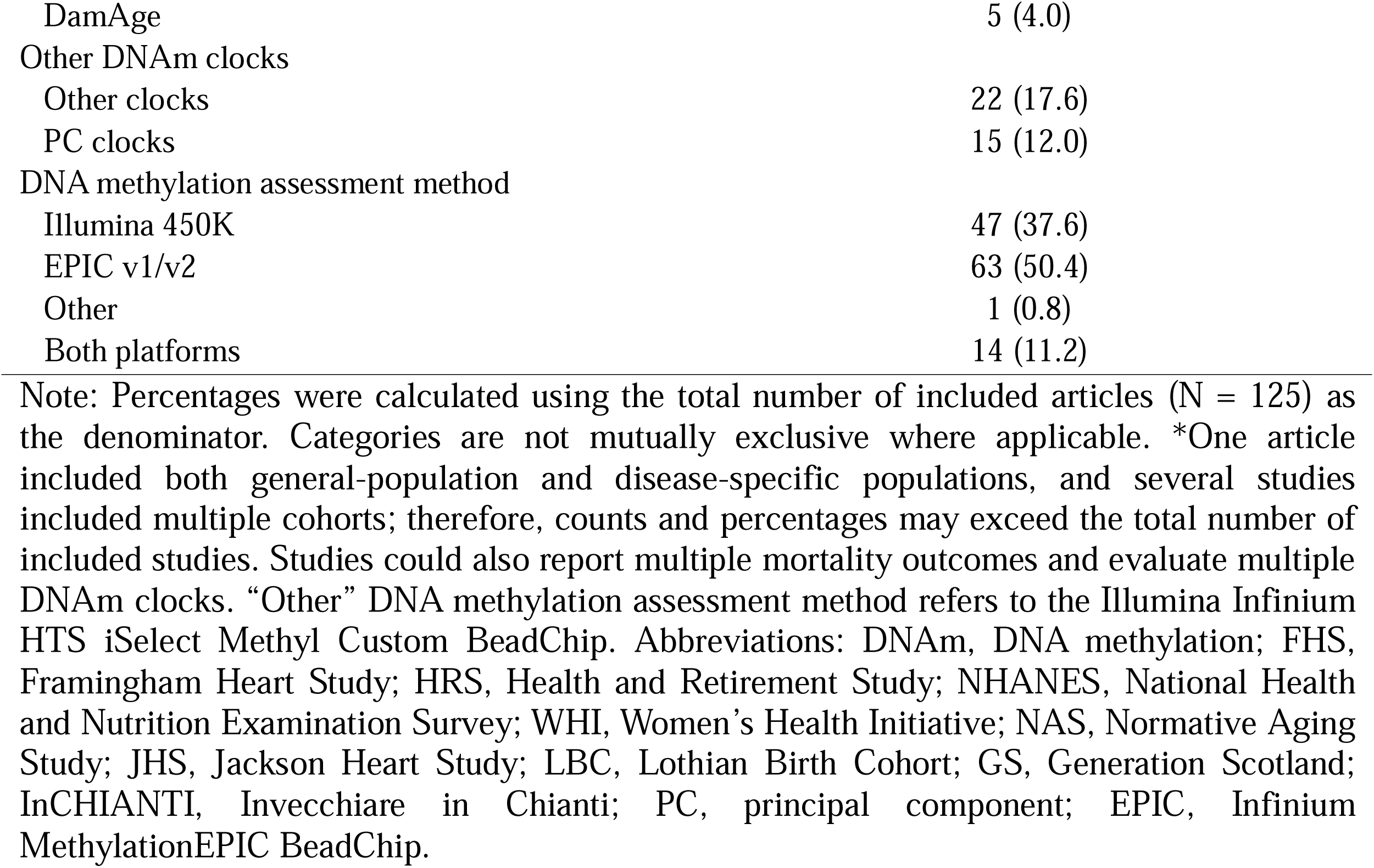
Summary of characteristics of included articles.

| Characteristics | Articles, n (%) |
| --- | --- |
| Total | 125 (100) |
| Outcomes |  |
| All-cause mortality | 116 (92.8)* |
| Cardiovascular mortality | 23 (18.4) |
| Cancer mortality | 22 (17.6) |
| Health status |  |
| General population | 97 (77.6) |
| Specific condition or disease | 29 (23.2) |
| Cohorts |  |
| FHS | 18 (14.4) |
| HRS | 17 (13.6) |
| NHANES | 17 (13.6) |
| WHI | 11 (8.8) |
| NAS | 11 (8.8) |
| JHS | 7 (5.6) |
| LBC1936 | 12 (9.6) |
| LBC1921 | 11 (8.8) |
| GS | 6 (4.8) |
| InCHIATI | 7 (5.6) |
| Others | 71 (56.8) |
| Cohort age |  |
| Mean/median age $\geq 65$ years | 52 (41.6) |
| Mean/median age $< 65$ years | 49 (39.2) |
| Both age groups or not reported | 24 (19.2) |
| First-generation DNAm clocks |  |
| Horvath1 | 74 (59.2) |
| Hannum | 68 (54.4) |
| Horvath2 | 16 (12.8) |
| Lin | 12 (9.6) |
| Weidner | 9 (7.2) |
| Vidal-Bralo | 8 (6.4) |
| Zhang2019 | 12 (9.6) |
| Second-generation DNAm clocks |  |
| PhenoAge | 74 (59.2) |
| GrimAge | 63 (50.4) |
| GrimAge2 | 23 (18.4) |
| FitAge | 3 (2.4) |
| ZhangMRscore | 13 (10.4) |
| DNAmTL | 15 (12.0) |
| Third-generation DNAm clocks |  |
| DunedinPoAm | 26 (20.8) |
| DunedinPACE | 35 (28.0) |
| Causal-generation clocks |  |
| AdaptAge | 5 (4.0) |
| CausAge | 4 (3.2) |
| DamAge | 5 (4.0) |
| Other DNAm clocks |  |
| Other clocks | 22 (17.6) |
| PC clocks | 15 (12.0) |
| DNA methylation assessment method |  |
| Illumina 450K | 47 (37.6) |
| EPIC v1/v2 | 63 (50.4) |
| Other | 1 (0.8) |
| Both platforms | 14 (11.2) |

Note: Percentages were calculated using the total number of included articles (N = 125) as the denominator. Categories are not mutually exclusive where applicable. \*One article included both general-population and disease-specific populations, and several studies included multiple cohorts; therefore, counts and percentages may exceed the total number of included studies. Studies could also report multiple mortality outcomes and evaluate multiple DNAm clocks. “Other” DNA methylation assessment method refers to the Illumina Infinium HTS iSelect Methyl Custom BeadChip. Abbreviations: DNAm, DNA methylation; FHS, Framingham Heart Study; HRS, Health and Retirement Study; NHANES, National Health and Nutrition Examination Survey; WHI, Women’s Health Initiative; NAS, Normative Aging Study; JHS, Jackson Heart Study; LBC, Lothian Birth Cohort; GS, Generation Scotland; InCHIANTI, Invecchiare in Chianti; PC, principal component; EPIC, Infinium MethylationEPIC BeadChip.

First-generation DNAm clocks included Horvath1 (n=74), Hannum (n=68), Horvarth2 (n=16), Lin (n=12), Weidner (n=9), Zhang2019 (n=10), Vidal-Bralo (n=9), 7CpG clock (n=2),^96,117^ 27CpGWKDE (Weighted 2D-kernel density estimations) (n=1)^29^, while second-generation clocks comprised of PhenoAge (n=74), GrimAge (n=63), GrimAge2 (n=23), ZhangMRscore (n=13), DNAmTL (n=15), FitAge (n=3), EpiTOC (mitotic age) (n=1),^104^ 59-CpG clock (single-stage mortality clock) (n=1),^6^ MetabolicAge (n = 1),^32^ MetaboHealth (n=1),^33^ intrinsic capacity clock (DNAm IC) (n=1),^34^ socioeconomic status clocks (aSES-BIO and cSES-BIO) (n=1)^35^ and bAge (n=1).^36,106^ Additionally, second-generation clocks also included two saliva/buccal based DNAm clocks, buccal clock (CheekAge) (n=1)^37^ and saliva clock (InflammAge) (n=1).^38^ Newer clocks included PhysAge,^53^ OMICmAge,^92^ DNAmEMRAge,^92^ InflaMeS,^64^ SenMortalityAge,^47^ SenChronoAge,^47^ SenCultureAge,^47^ Bocklandt,^63^ Garagani,^63^ Universal pan-mammalian,^144^ and DeepStrataAge.^109^ PC clocks (PCHorvath1, PCHorvath2, PCHannum, PCPhenoAge, PCGrimAge, PCDNAmTL) were reported in 15 studies.^11,54,56,58,61,90–92,102,106,117,119,135,137,144^ Third-generation clocks included DunedinPace (n=35) and DunedinPoAm (n=26). Fourth-generation clocks incorporated causal clocks (AdaptAge, DamAge and CausAge) (n=5, 5 and 4, respectively) and organ system-specific clock, SystemsAge (n=1).^11^

All-cause mortality was the most assessed outcome (n=116), followed by cardiovascular mortality (n=23) and cancer mortality (n=22) (Table 1). Detailed study characteristics and effect sizes are provided (**Supplementary Table 2-3**). Overall, 106 articles were rated as high quality, 16 as moderate quality, and three as low quality (**Figure 2, Supplementary Table 4**).

**Fig. 2:**
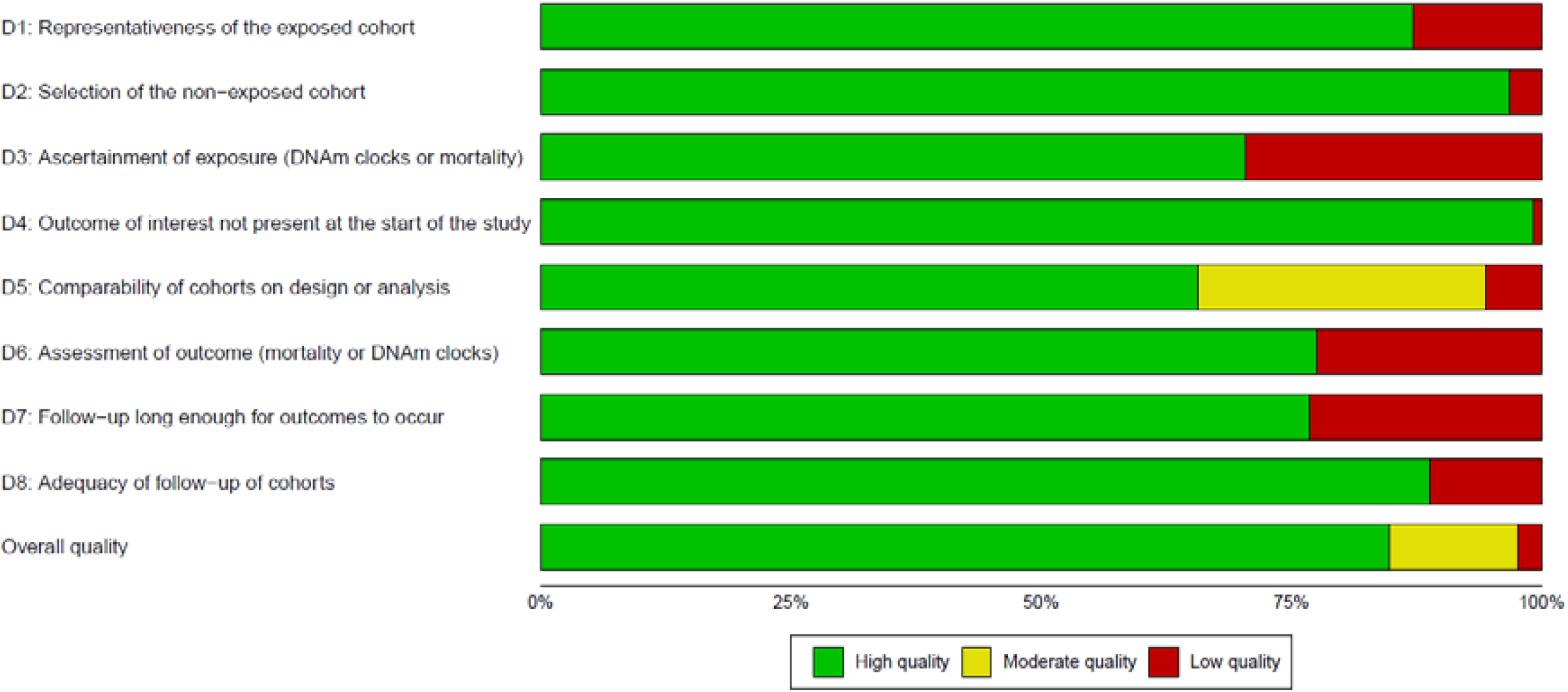
Methodological quality of the 125 included studies. Stacked bars show the percentage of the 125 included cohort studies rated high quality (green), moderate quality (yellow) or low quality (red) on each item of the Newcastle–Ottawa Scale for cohort studies, as adapted for this review, and on overall quality.

### Summary of included articles

For all-cause mortality, 92 articles in the general populations (n=343 381 participants) and 23 articles (n=24 784 participants) in diseased-specific populations, and one article reporting both populations (general population n = 2805, disease-specific population n= 582) were included in the systematic review. In the general population, first-generation clocks, Horvath1 (38%, 17/45 unique cohorts) and Horvath2 (27%, 4/15), were not significantly associated with all-cause mortality, while Hannum (65%, 28/43), Zhang2019 (80%, 4/5) were significantly and positively associated with all-cause mortality (**Figure 3A**). Second and third-generation clocks, including PhenoAge (91%, 31/34), GrimAge (100%, 37/37), GrimAge2 (96%, 22/23), ZhangMRscore (83%, 15/18), DunedinPoAm (77%, 10/13), and DunedinPace (96%, 25/26), consistently demonstrated positive and statistically significant associations, while DNAmTL (70%, 14/20) showed a significant negative association. Newer generation of clocks including DamAge (80%, 4/5), CausAge (80%, 4/5) and SystemsAge (100%, 7/7) were positively significantly associated with all-cause mortality. The bAge, DNAm IC, FitAge, DNAm Metabolic, MetabolicHealth, 59 CpG, aSES-BIO, cSES-BIO, InflammAge, and CheekAge^37^, showed significant positive associations, but were reported in a limited number of cohorts (n ≤5). Similar patterns, particularly for second-generation clocks, were observed in diseased-specific populations across different disease cohorts.

**Fig. 3:**
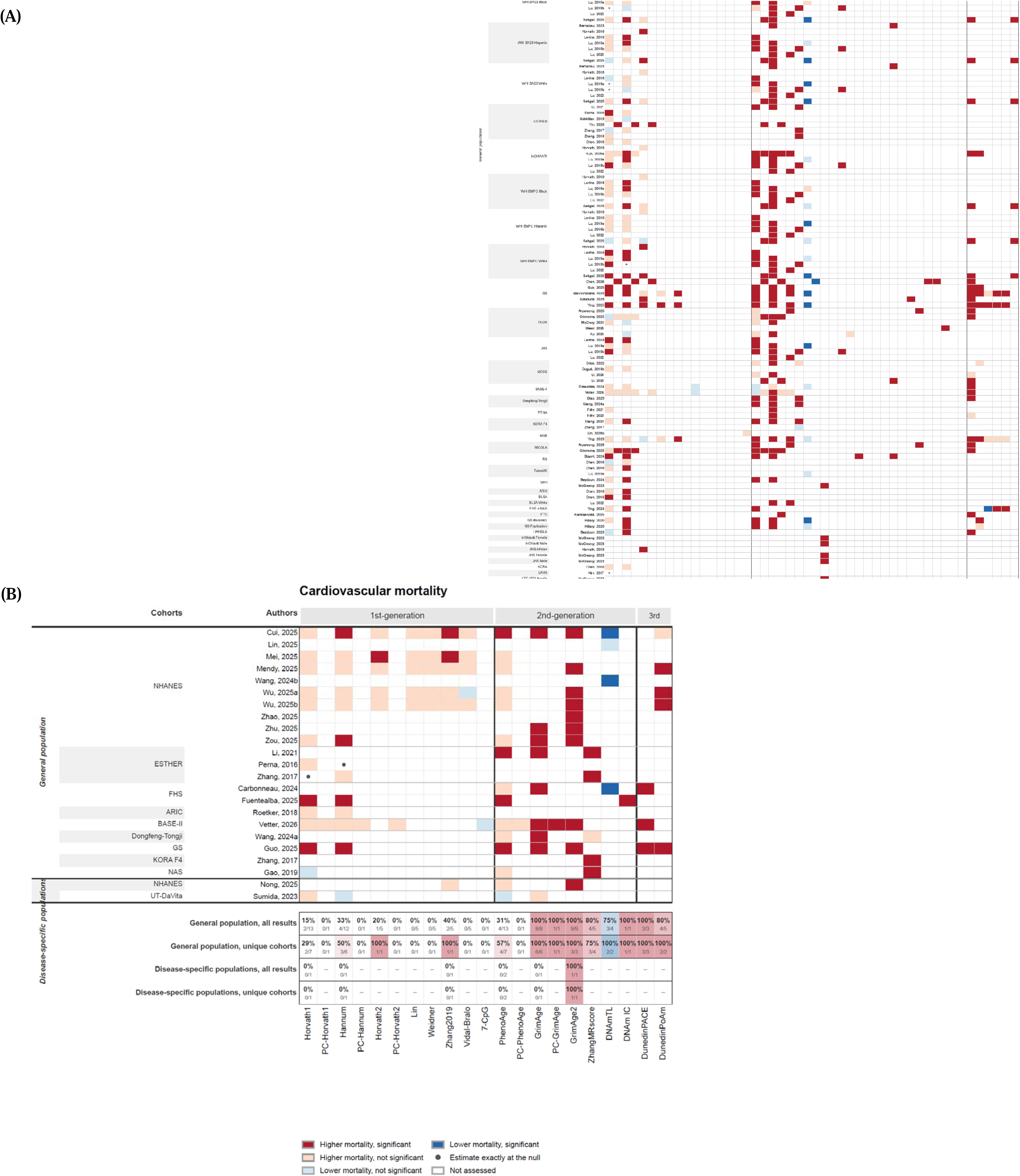
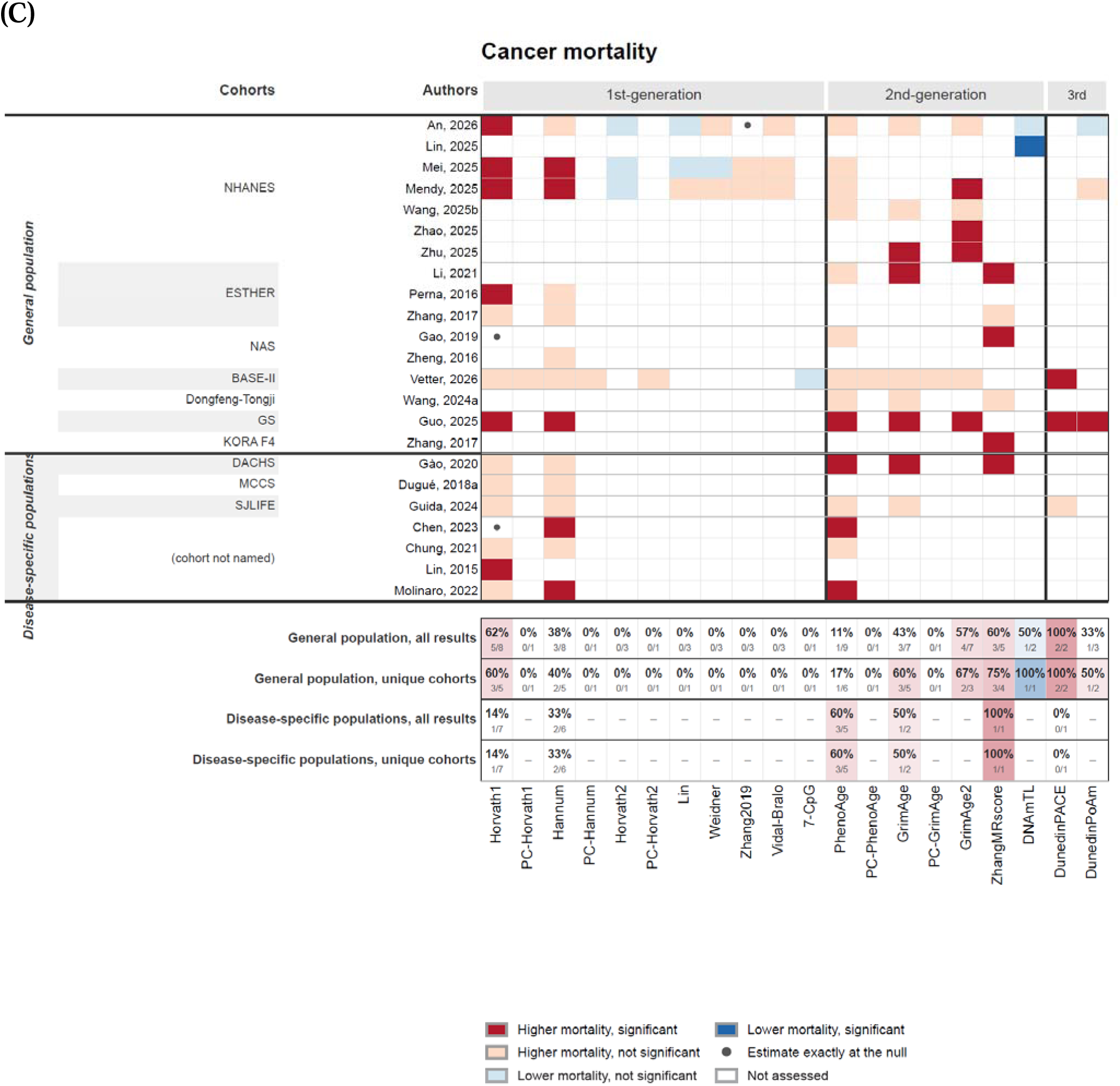
Heatmaps of the associations between DNA methylation clocks and mortality across all included studies. a–c, Heatmaps of all-cause mortality (a), cardiovascular mortality (b) and cancer mortality (c). Rows are individual studies (first author and year) with the contributing cohort or sub-cohort named alongside, blocked into general populations and disease-specific populations; columns are DNA methylation (DNAm) clocks, ordered and bracketed by clock generation. Cell colour encodes the direction and statistical significance of the fully adjusted association as reported by the original study: significantly higher mortality (dark red), higher mortality but not significant (pale red), lower mortality but not significant (pale blue), significantly lower mortality (dark blue), an estimate exactly at the null (dot), significance not determinable from the report (mid grey), clock assessed but no estimate reported (light grey) and clock not assessed (white). Beneath each heat map, four summary rows give the percentage (and count) of determinable estimates that were significant in the expected direction, a higher hazard for all clocks except DNAmTL, PCDNAmTL, AdaptAge and DNAmIC for which a lower hazard is expected, computed over all results and over unique cohorts (a cohort counts once, and counts as significant if any report on it was), separately for general and disease-specific populations.

For cardiovascular mortality, 21 studies in the general population (n=58 628 participants)^13,30,34,44,70,72,73,77–82,84,100,108,110,114,115,117,118^ and two (n = 514 participants)^74,142^ in a diseased population were included (**Figure 3B**). In the general population, Horvath1 was not associated with cardiovascular mortality across cohorts (29%, 2/7), whereas Hannum (50%, 3/6) showed inconsistent associations. Hovarth2 (100%, 1/1), Zhang2019 (100%, 1/1), PhenoAge (57%, 4/7), GrimAge (100%, 6/6), GrimAge2 (100%, 3/3), ZhangMRscore (75%, 3/4), DNAmTL (100%, 2/2), DunedinPoAm (100%, 2/2), and DunedinPace (100%, 3/3) were significantly associated with higher cardiovascular mortality, although each was reported in five or fewer unique cohorts. Among disease-population, only GrimAge2 was associated with cardiovascular mortality in people with diabetes^74^.

For cancer mortality, 15 studies in the general population (n=44 449 participants) ^30,68,72,73,77,81,82,84,87,100,108,110,114,117,118^ and 7 studies in a diseased population (cancer, cancer survivors, or glioma) (n = 9 000 individuals)^122,124–129^ were included in the systematic review (**Figure 3C**). In the general population, Hovarth1 was significantly associated with cancer mortality (60%, 3/5), whereas Hannum (40%, 2/5), PhenoAge (17%, 1/6) and other first-generation clocks were not. GrimAge (60%, 3/5), GrimAge2 (67%, 2/3), ZhangMRscore (75%, 3/4) were significantly associated with cancer mortality. DNAmTL and DunedinPace were significantly associated with cancer mortality in 2 or less unique cohorts. In disease-specific populations, GrimAge (50%, 1/2) and ZhangMRscore (100%, 1/1) were associated with cancer mortality in a population with colorectal cancer and cancer survivors.^122,127^

### Meta-analyses

Analyses were restricted to general population cohorts and not performed in the disease-specific populations due to clinical heterogeneity. Fifty-three articles were included in the meta-analyses across DNAm clocks for all-cause, cardiovascular and cancer mortality.

For all-cause mortality, all DNAm clocks except CausAge, EpiTOC, AdaptAge and Zhang2019 showed significant associations in minimally adjusted models (**Figure S2A**, **Figure S3A** and **Supplementary Table 6**). In fully adjusted models, the largest pooled HR was observed for PCGrimAge EAA (8 cohorts, n = 28 320, HR per SD: 2.10 [95%CI 1.15, 3.82], I²=98.6%), followed by GrimAge2 EAA (22 cohorts, n=40 258, HR per SD: 1.65 [95% CI 1.53, 1.77], I²=77.5%), bAge (6 cohorts, HR per SD: 1.52 [95%CI: 1.38, 1.68], I²=54.2%) and GrimAge (31 cohorts, n=53 642, HR per SD: 1.46 [95% CI 1.38, 1.55], HR per 1-year: 1.09 [95% CI 1.08, 1.11], I²=84.1%) (**Figure 4A, Figure S1A, Supplementary Table 5 & 10**). Technical refined clocks (PCGrimAge) and clocks with enhancement with additional surrogates (GrimAge2 and bAge) had larger pooled HRs than its predecessor (GrimAge). Funnel plots indicated asymmetry for GrimAge and GrimAge2, which was supported by Egger’s tests (p=0.0016 and p=0.0105, respectively), suggesting potential small-study effects (**Figures S4-6 and S7, Supplementary Table 5-6**). Small-study effects could not be assessed for PCGrimAge or bAge, which were reported in fewer than ten cohorts (8 and 6, respectively). HR expressed per 1-year were reported in **Supplementary Tables 5-6**.

**Fig. 4:**
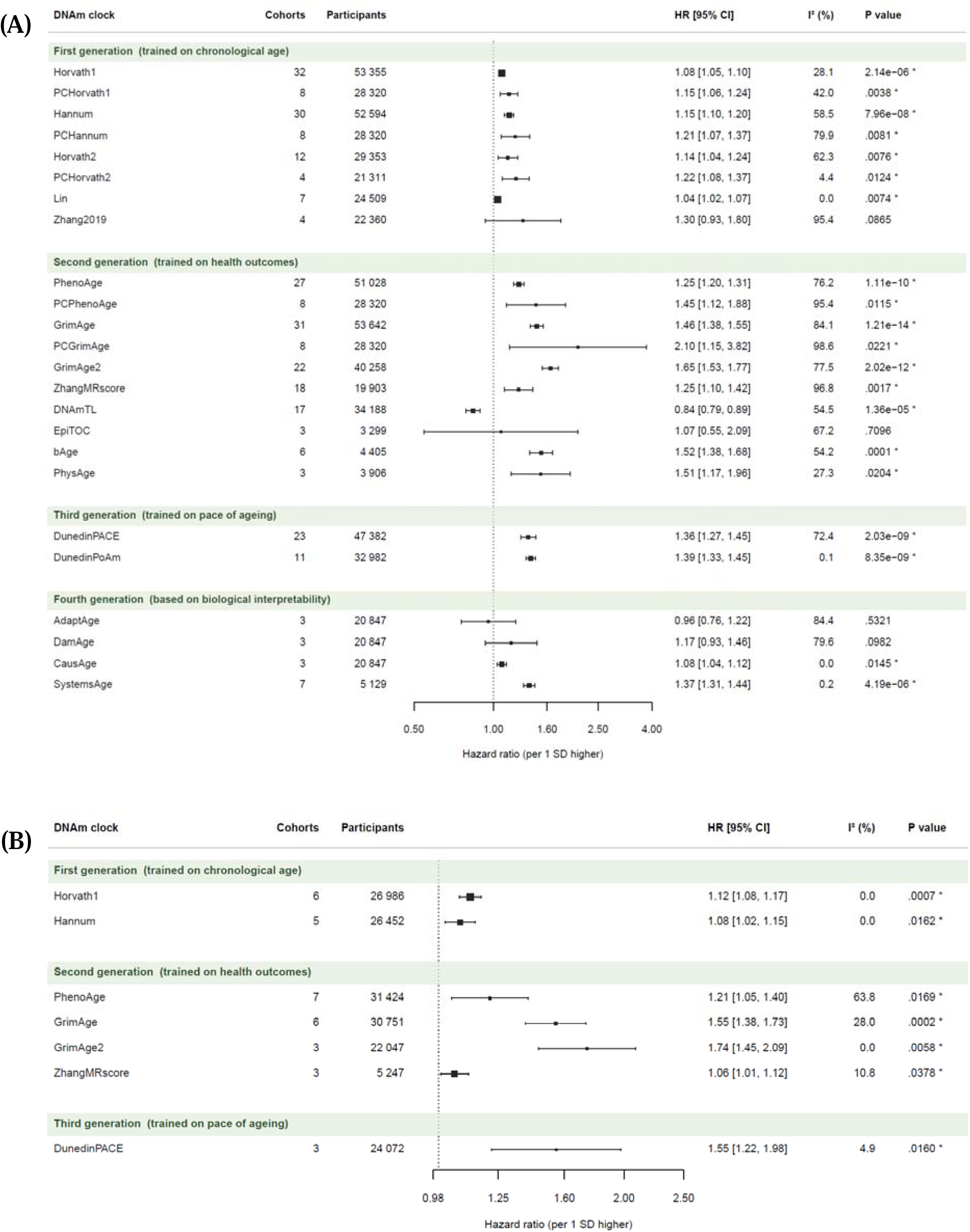
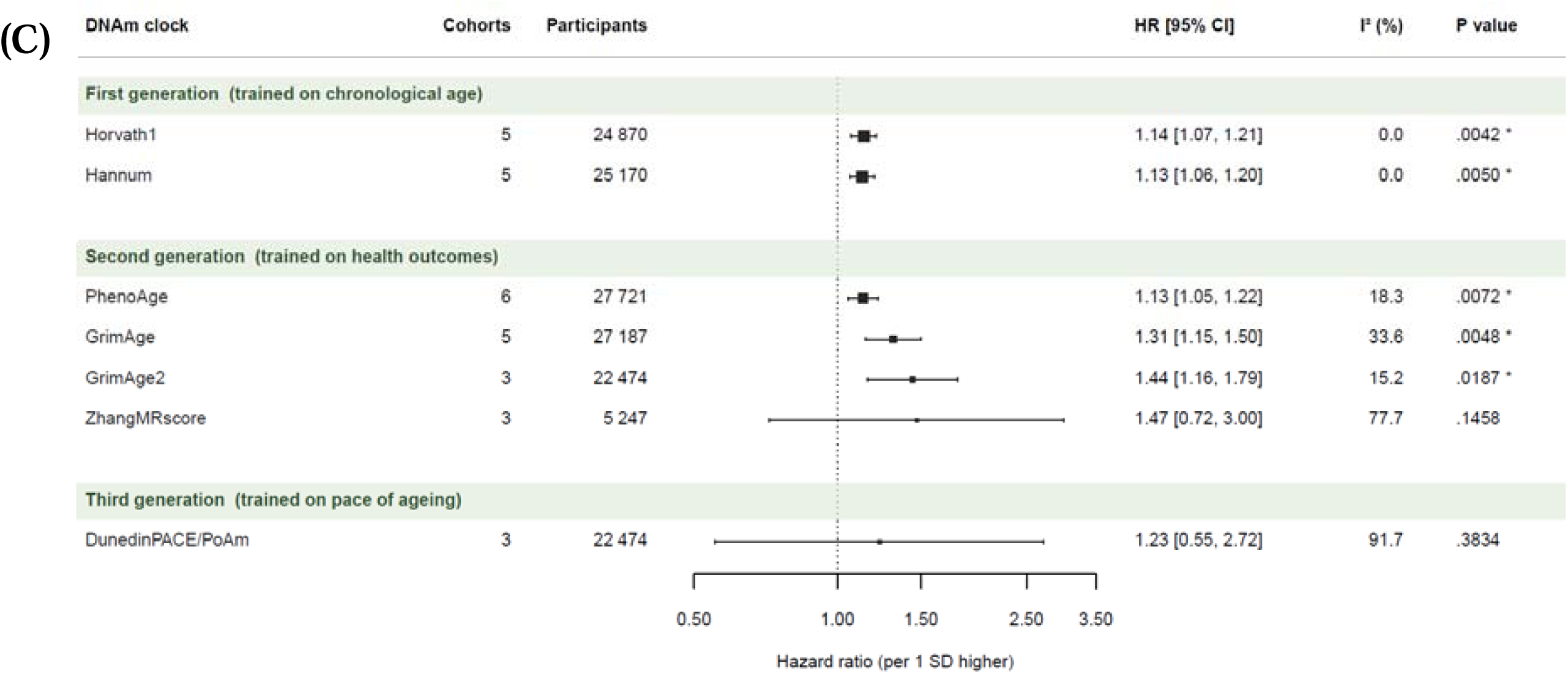
Meta-analytic association between DNA methylation clock EAA and mortality in general-population cohorts. a–c, Random-effects meta-analyses of fully adjusted models for all-cause mortality (a), cardiovascular mortality (b) and cancer mortality (c). Each row is one DNA methylation (DNAm) clock with at least three independent cohorts; rows are blocked by clock generation. Squares denote the pooled hazard ratio (HR) per 1 s.d. higher EAA, with square area proportional to the precision of the pooled estimate, and horizontal error bars denote the 95% confidence interval (CI); the x axis is on a logarithmic scale and the vertical reference line marks the null (HR = 1). Columns to the right give the number of contributing cohorts, the number of participants (n), the pooled HR [95% CI], the I² statistic and the two-sided P value for the pooled estimate; an asterisk marks P < 0.05. Minimally adjusted models, per-1-year estimates and cohort-level forest plots are given in Supplementary Figs. 2 and 3 and Supplementary Table 6.

Sensitivity analysis restricted to cohorts rated high quality resulted in no statistically significant attenuation (**Supplementary Tables 7 and 8**). Excluding effect estimates whose conversion to a per-SD scale relied on an imputed clock standard deviation left the pooled HRs largely unchanged where pooling remained possible (**Supplementary Table 9**). These indicate the robustness of the main findings.

For cardiovascular mortality, all DNAm clocks were significantly associated with mortality in minimally adjusted models (**Figure S2B, Figure S3B and Supplementary Table 6**). In fully adjusted models, the largest pooled HR was observed for GrimAge2 EAA (HR per SD, 1.74 [1.45, 2.09]; I²=0%), followed by DunedinPACE (HR per SD, 1.55 [95% CI 1.22, 1.98]; I²=4.9%) and GrimAge EAA (HR per SD, 1.55 [1.38, 1.73]; HR per 1-year: 1.10 [95% CI 1.07, 1.13], I²=28.0%) (**Figure 4B, Figure S1B and Supplementary Table 5**). Sensitivity analysis restricted to high-quality cohorts left every cardiovascular estimate unchanged, as all contributing cohorts were rated high quality (**Supplementary Tables 7 and 8**). Excluding effect estimates that relied on an imputed clock standard deviation left the pooled HRs largely unchanged where pooling remained possible (**Supplementary Table 9**).

For cancer mortality, all DNAm clocks were significantly associated with mortality in minimally adjusted models, except for ZhangMRscore (**Figure S2C, Figure S3C and Supplementary Table 6**). In fully adjusted models, the largest pooled HR was observed for GrimAge2 EAA (HR per SD, 1.44 [1.16, 1.79]; I²=15.2%), followed by GrimAge EAA (5 cohorts, n=27 187, HR per SD: 1.31 [95% CI 1.15, 1.50], HR per 1-year: 1.06 [95% CI 1.03, 1.10], I²=33.6%), whereas DunedinPACE was no longer associated with mortality (p=0.383) (**Figure 4C, Figure S1C and Supplementary Table 5**). Sensitivity analysis restricted to high-quality cohorts left every cancer estimate unchanged, as all contributing cohorts were rated high quality (**Supplementary Tables 7 and 8**). Excluding effect estimates that relied on an imputed clock standard deviation left the pooled HRs largely unchanged where pooling remained possible (**Supplementary Table 9**).

### Improved performance beyond GrimAge

On a per-SD scale, GrimAge2 consistently yielded larger pooled HRs than GrimAge across all outcomes, adjustment models and sensitivity analyses. For fully adjusted all-cause mortality, the CIs overlapped only marginally (GrimAge2 HR 1.65 [95% CI 1.53, 1.77] vs GrimAge 1.46 [1.38, 1.55]; **Figure 4A**).

In six cohorts (FHS, LBC1921, LBC1936, WHI BA23-Black, WHI BA23-Hispanic, WHI BA23-White) bAge (another enhanced version of GrimAge) yielded larger HR and narrower 95% CI for all-cause mortality (HR per SD: 1.52 [1.38, 1.68]) compared to GrimAge (HR per SD: 1.47 [1.30, 1.67] (**Figure 5, Supplementary Table 11-12**). These differences should be interpreted as indicative evidence rather than as evidence of statistical superiority given the overlapping CI.

**Fig. 5:**
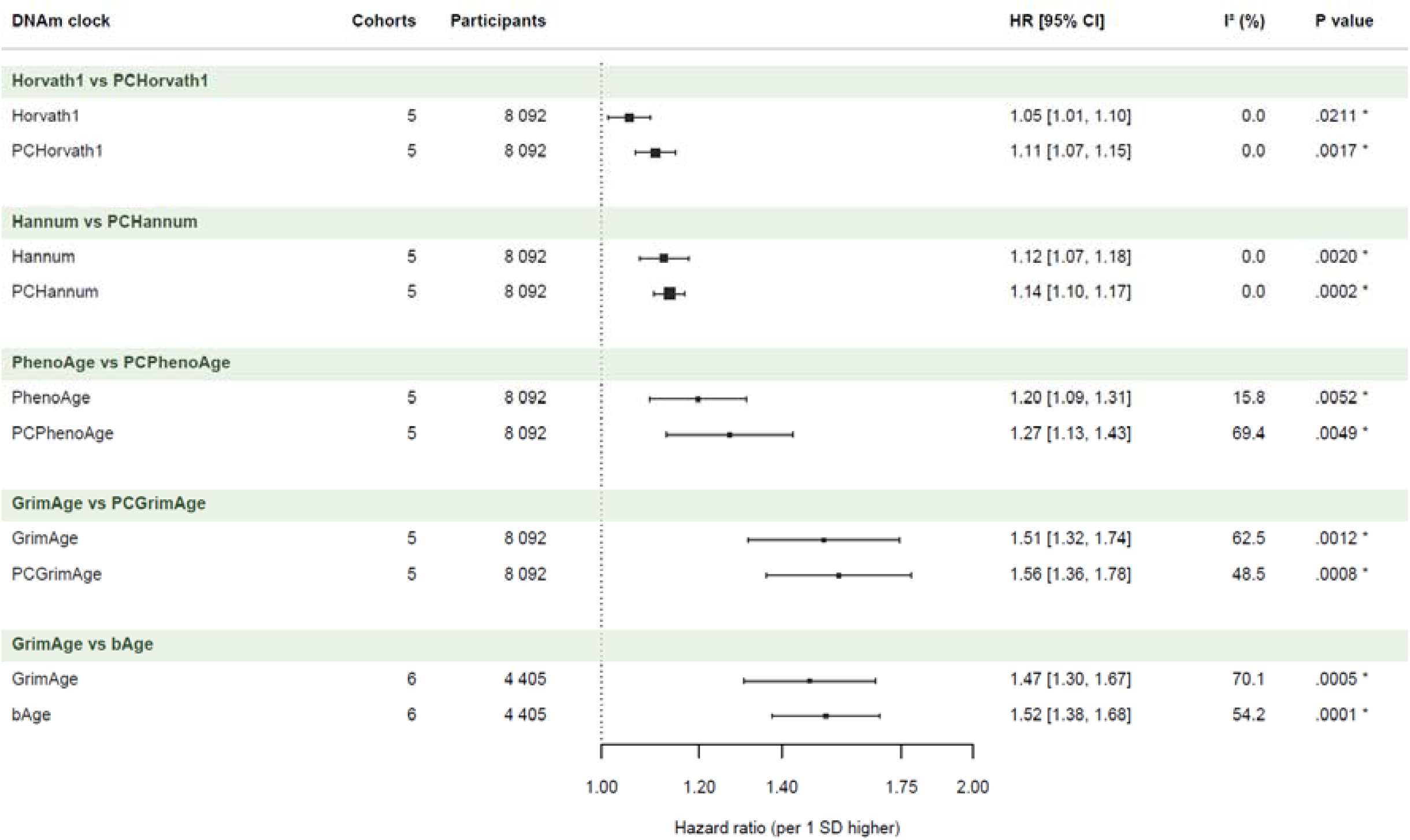
Head-to-head comparison of original DNA methylation clocks and their enhanced counterparts within matched cohort in the same articles. Random-effects meta-analyses restricted, within each block, to the cohorts that reported both members of the pair: Horvath1 versus PCHorvath1, Hannum versus PCHannum, PhenoAge versus PCPhenoAge and GrimAge versus PCGrimAge (five cohorts, n = 8,092 participants each), and GrimAge versus bAge (six cohorts, n = 4,405 participants). Squares denote the pooled hazard ratio (HR) for all-cause mortality per 1 s.d. higher EAA, with square area proportional to the precision of the pooled estimate, and horizontal error bars denote the 95% confidence interval (CI); the x axis is on a logarithmic scale and the vertical reference line marks the null (HR = 1). Columns to the right give the number of contributing cohorts, the number of participants (n), the pooled HR [95% CI], the I² statistic and the two-sided P value; an asterisk marks P < 0.05. Pooling followed the same model as in Fig. 4.

### Performance of PC clocks

In five cohorts (BASE-II, InChianti, TILDA, NICOLA, and HRS) that evaluated both the original clocks and their PC-enhanced counterparts, the PC clocks yielded consistently higher hazard ratios for mortality risk (**Figure 5, Supplementary Table 11-12**). One SD higher PCGrimAge was associated with 56% higher all-cause mortality (HR per SD = 1.56 [1.36, 1.78]), compared with 51% for GrimAge (HR per SD = 1.51 [1.32, 1.74]). The same direction was observed for the Horvath, Hannum and PhenoAge clocks (PCHorvath1: 1.11 [1.07, 1.15] vs Horvath1 1.05 [1.01, 1.10]; PCHannum: 1.14 [1.10, 1.17] vs Hannum: 1.12 [1.07, 1.18]; PCPhenoAge: 1.27 [1.13, 1.43] vs PhenoAge: 1.20 [1.09, 1.31]). These differences should be interpreted as indicative evidence rather than as evidence of statistical superiority given the overlapping CI.

## Discussion

### Summary of evidence

This systematic review found that DNAm clocks are predictive of all-cause, cardiovascular, and cancer mortality across both general and disease-specific populations. In meta-analyses, GrimAge EAA was associated to all-cause, cardiovascular, and cancer mortality. Technical refinements, including PC correction and iterative enhancement of existing clocks (bAge and GrimAge2 refined from GrimAge), further strengthened predictive performance by reducing technical noise and incorporating richer biological information.^7,33,36,43^ These findings were not reported in previous systematic reviews of DNAm clocks and mortality.^14–16^

### GrimAge and traditional biomarkers

Most DNAm clocks showed significant predictive ability for all-cause mortality, suggesting their potential as biomarkers of ageing. Additionally, GrimAge EAA showed strong and consistent predictive ability with all-cause mortality, with effect sizes that were comparable in magnitude to, or larger than, those reported for established cardiometabolic, inflammatory, and fitness-related biomarkers.^146,147^ In a nationwide cohort of 12.4 million Korean adults, a 1-SD higher fasting glucose (27.5 mg/dL) was associated with a 21% higher risk of all-cause mortality.^146^ In 411 506 UK Biobank participants, a 1-SD (4.31mg/ml) higher C-Reactive Protein was associated with a 13% higher risk of all-cause mortality.^147^ Cardiorespiratory fitness showed protective effects: in 70 913 UK Biobank participants, a 1-SD higher cardiorespiratory fitness was associated with a 23% lower mortality risk^148^, and across 3.8 million adults from 35 cohorts, each higher metabolic equivalent of task (3.5 mL/kg/min) was associated with a 14% lower mortality risk, corresponding to a 17–23% lower mortality risk per 1-SD (4.31-6.06 mL/kg/min) higher in VO₂max. In contrast, a 1-SD higher GrimAge EAA (3–5 years) in this meta-analysis was associated with a 33–61% higher risk of all-cause mortality. Therewith, GrimAge potentially captures stronger mortality-relevant biological ageing signals, although direct comparisons within the same cohorts are required to establish incremental predictive value over traditional biomarkers. By demonstrating strong and consistent predictive ability of DNAm clocks for mortality across multiple cohorts and adjustment models, this review provides empirical support for their potential candidacy for clinical use, although causality remains unproven.^3^

### Technical refinement

Clocks with technical refinement consistently outperform the original clocks. This aligns with previous studies suggesting that PC analysis reduces technical noise, enhances longitudinal stability, offers better calibration in heterogeneous multi-cohort settings and greater utility for epidemiological and translational work.^43,149–151^ Enhancement of existing clocks is well demonstrated by the progression from GrimAge to GrimAge 2,^7^ GrimAge with additional DNAm proxies of metabolites from MetaboHealth,^33^ as well as GrimAge to bAge (age, GrimAge DNAmPackYears, 5/7 GrimAge protein proxies (B2M, cystatin C, GDF15, PAI1, TIMP1), and 28/109 protein EpiScores^152^).^36^ These enhancements consistently yield stronger all-cause mortality prediction, indicating that incremental enhancement, rather than entirely new clocks, can meaningfully improve predictive performance.^7,33,36^ The improved performance of PC-clocks and enhanced models indicates that future DNAm clocks should incorporate PCA-based noise correction and systematic model refinement during the training phase.

### Saliva-based clocks

A few non-blood-based-DNAm clocks were included in this study and showed predictive ability for mortality. CheekAge demonstrated weaker mortality prediction than PhenoAge but stronger performance than first-generation clocks, while exhibiting the highest test–retest reliability in buccal tissue and high accuracy in saliva samples.^37^ In addition, InflammAge, a saliva-based clock outperformed Horvath2 while performing in the range of PhenoAge EAA.^38^ Application of blood-based DNAm clocks in saliva samples showed the greatest variability across DNAm arrays compared to buffy coat and peripheral blood mononuclear cell, largely due to heterogeneous cell-type composition and higher sampling and technical variations in saliva.^149,153,154^ Therefore, instead of applying blood-based DNAm clocks on buccal or salivary samples, CheekAge and InflammAge may represent more suitable candidates for these samples.^37^

### Strengths and limitations

The strengths of this review include the incorporation of DNAm clocks across four generations, which have been underrepresented in previous reviews. Limitations include substantial between-study heterogeneity and publication bias. Publication bias may inflate pooled associations through underrepresentation of null findings; therefore, estimates for clocks evaluated in few cohorts should be interpreted cautiously. However, sensitivity analyses and Hartung–Knapp adjustment supported the robustness of the main findings, despite the substantial heterogeneity. Given the predominance of retrospective cohort studies, although sensitivity analyses restricted to high-quality studies yielded similar results, NOS-based quality ratings should be interpreted cautiously, and the meta-analysis outcomes from this study are unable to conclude causal inference. Additionally, comparisons between DNAm clocks based on meta-p-values should be interpreted cautiously, as these are influenced by both the number and size of included cohorts.

### Future direction

To further understand the clinical applications of DNAm clocks, clinical trials including pharmacologic or lifestyle-based interventions that monitor DNAm clocks as outcomes will be essential for establishing their responsiveness and evaluating whether they meet criteria for clinical surrogate endpoints. Such trials will also generate the data needed to develop new DNAm-derived surrogates that are directly calibrated to intervention-induced biological change. Integrating these clinically responsive surrogates with multi-omic DNAm proxies, including proteomic EpiScores^152^ (as in bAge^36^) or metabolomic components (as in metabolic-enhanced GrimAge),^33^ may further enhance both predictive accuracy and mechanistic insight. Incorporating PCA during clock construction reduces technical heterogeneity and improves cross-cohort robustness, and should therefore be standard practice in future clock development and clinical application.^43,149–151^ For studies using saliva or buccal samples, CheekAge should be adopted as a benchmark due to its superior reliability in these tissues.^37^

### Conclusions

In conclusion, DNAm EAAs are predictive of higher all-cause, cardiovascular and cancer mortality risk. GrimAge is the predictor with the most robust association for mortality prediction, and continued refinement through models such as GrimAge2, bAge, and metabolite-enhanced GrimAge illustrates the value of iterative improvement. Together, methodological refinement (PC clocks) and enhancement of existing clocks offer a clear pathway toward clinical utility in precision geromedicine. Critical gaps remain, including clarification of causal mechanisms, demonstration of responsiveness to interventions, and evaluation of incremental value over existing clinical predictors within the same cohorts using measures such as changes in AUC. With concerted efforts in standardisation, multi-omic integration, and intervention trials, DNAm clocks hold high potential to move from epidemiological insight to actionable biomarkers of ageing.

## What is already known on this topic

- DNAm clocks have been associated with all-cause mortality in multiple cohorts.
- Prior syntheses suggest that second-generation mortality-trained clocks (eg, GrimAge) outperform first-generation chronological-age clocks (Horvath, Hannum) for mortality prediction.
- Existing systematic reviews and meta-analyses have evaluated only a subset of clocks (mainly Horvath, Hannum, PhenoAge, and GrimAge) and do not reflect the rapid expansion of newer clocks developed after GrimAge.

## What this study adds

- This updated systematic review synthesized evidence from 125 studies (382 307 participants) evaluating 48 DNAm clocks, showing that higher DNAm age measures are associated with higher risks of all-cause, cardiovascular, and cancer mortality across both general and disease-specific adult populations.
- In meta-analyses of general populations, GrimAge showed among the largest pooled associations and consistent mortality associations: each 1-year higher GrimAge EAA was associated with approximately 9% higher all-cause, 10% higher cardiovascular, and 6% higher cancer mortality risk.

The review substantially expands previous evidence by evaluating newer clocks, including GrimAge2, DunedinPACE, DNAmTL, SystemsAge, bAge, causal clocks, and PC-corrected clocks. Technically refined clocks, particularly GrimAge2 and PC-enhanced clocks, generally showed stronger mortality associations than their original counterparts, supporting further evaluation of their incremental predictive value, intervention responsiveness, and clinical utility.

## Data availability

All data generated or analysed in this study are included in the Supplementary Material. All extracted effect sizes used for the meta-analyses, together with the corresponding graphical summaries, are provided in the Supplementary Material.

## Ethical approval

Not required

## Acknowledgments

This Article is funded by an unrestricted grant (startup grant; A-0009805-00-00) by the National University of Singapore to ABM. We thank Senior Librarian, Loh Mee Lan, from NUS Libraries National University of Singapore, for assistance in constructing the initial search strategy. This systematic review is part of the Global Epigenetic Age Consortium, dedicated to synthesising evidence on the association of DNAm clocks with clinically relevant outcomes including age-related diseases, frailty and mortality and therewith advancing the potential clinical translation of DNAm clocks for geroscience and precision geromedicine.

## Contributors

JHT and WW designed the screening strategy and independently conducted the abstract and full-text screening. JHT, DB, JD, SC and AB independently extracted data from eligible studies and drafted the initial manuscript, after which JHT carried out the meta-analysis and interpreted the results. WW, DB and ABM proofread the manuscript, providing critical feedback. WW and ABM supervised the entire process, offering methodological guidance and overseeing manuscript revisions. All authors approved the final version of the manuscript. JHT, DB, JD, SC, AB, WW, and ABM had full access to the data and verified its accuracy.

