## Supplementary Text 1 for "Biological age measured by DNA methylation clocks and mortality: A systematic review and meta-analysis"

### Supplementary Text 1. Details of method section

**Search strategy and selection criteria**

The search date was set from 1 January 2011 until 15th April 2026 (the first search was conducted on 10th September 2024, and the updated search on 6th June 2025 and 15th April 2026). During the first literature search, a large number of relevant studies was identified, making it impractical to summarise all outcomes in a single manuscript. Therefore, in the present paper, the scope was narrowed to two important age-related outcomes, epigenetic age and mortality. Moreover, the initial registration provided examples of age-related diseases and conditions for each physiological system. However, this list was not exhaustive.

**Article selection**

The inclusion criteria consisted of peer-reviewed population-based cohort articles written in English, including adult humans with a mean or median age of 18 years old and above, examining the associations between DNAm clocks and mortality including all-cause mortality, cardiovascular mortality and cancer mortality. Cohort studies including the general population and populations of specific diseases with a control group were included. A control group is essential because it provides a baseline to ensure mortality observed was not due to those of underlying conditions. Only articles that included outcomes or predictors specifically related to DNAm clocks, such as DNAm age, DNAm age acceleration (AgeAcce, also known as epigenetic age acceleration (EAA) or relative age acceleration) or DNAm age deviation (AgeDev, also known as absolute age acceleration) measurements, were considered. DNAm age refers to the actual estimate of a DNAm-based clock for a given sample that the clock is applied to. EAA refers to residuals of a linear regression of DNAm age against chronological age. AgeDev refers to the difference between DNAm age and an individual's chronological age. Higher DNAm age reflects higher biological age, whereas higher AgeDev and EAA indicate biological age exceeding chronological age.

**Data synthesis and Meta-analysis**

The meta-analyses were conducted with Hartung-Knapp adjustment to ensure more robust random-effects confidence intervals. *I²* representing the proportion of variability among study outcomes, along with the chi-squared test and its associated p-value, was calculated and presented in the forest plot. *I²* was defined as low (*I^2^* ≤25 %), moderate (*I^2^* >25 % and <75 %), or high (*I^2^* ≥75 %) heterogeneity. χ² (Cochran’s Q test) refers to the observed variability in study estimates exceeds that expected by sampling error alone; a low p-value suggests presence of heterogeneity. τ² (Tau-squared) refers to the estimates the absolute between-study variance in effect sizes, with larger values indicating greater heterogeneity; used in random-effects modelling to weight studies. For studies with missing demographic information (mean age, sex distribution, array platform, or clock standard deviations), values were imputed using data from other publications based on the same underlying cohort when available. When the standard deviation (SD) of the clock measure was not reported but alternative dispersion measures were provided, SD or standard error (SE) was derived using established conversion formulas: When standard error was not directly reported, we estimated them using:

- From 95% confidence intervals (95% CI):
  - SE = (95%CI upper limit – 95% CI lower limit)/3·92

When SD(X) or SD(Y) is not directly reported:

- From interquartile range (IQR):
  - SD(Y) = IQR/1·349
- From range:
  - SD(Y) = (maximum-minimum)/6
- From standard error:
  - SD(Y) = SE × √n
