## Supplementary Text 2 for "Biological age measured by DNA methylation clocks and mortality: A systematic review and meta-analysis"

### Supplementary Text 2. Newcastle-Ottawa Scale (NOS): Cohort studies

**Selection**

1) Representativeness of the exposed cohort
   a) truly representative of the average population: Age, gender distribution, country and kind of population are reported*
   b) Not representative based on factors mentioned above
   c) no description
2) Selection of the non-exposed cohort
   a) drawn from the same community as the exposed cohort *
   b) drawn from a different source
   c) no description of the derivation of the non-exposed cohort
3) Ascertainment of exposure: DNAm clocks or Mortality
a) Sample type: whole blood, PBMC, buffy coats or dried blood spot*
b) Sample type: peripheral blood or blood.
c) No description of the sample type.
4) Demonstration that the outcome of interest was not present at the start of the study
   a) yes *
   b) no

**Comparability (Max 2 stars, allow multiple selection)**

1) Comparability of cohorts on the basis of the design or analysis
   a) The study controls for the most important factor (Age and sex)*
   b) The study control for any additional factor (cell composition or lifestyle factors)*
   c) No controlling for any factors
   d) No description

**Outcome**

1) Assessment of outcome: Mortality or DNAm clocks
   a) Independent blind assessment (objective measures)*
   b) Record linkage (Clearly stated that information comes from national records, death certificates, or other medical or hospital records)*
   c) Exclusively through next of kin interview
   d) no description
2) Was follow-up long enough for outcomes to occur
   a) yes ≥ 5 years*
   b) no
3) Adequacy of follow up of cohorts
   a) complete follow up or small number lost - < 20% *
   b) Large number lost (≥ 20%)
   c) no statement
